# Metabolic associations with stroke, dementia, and imaging markers of cerebral small vessel disease: a comprehensive metabolomics study

**DOI:** 10.1101/2022.03.24.22272911

**Authors:** Eric L Harshfield, Hugh S Markus

## Abstract

**Background:** Cerebral small vessel disease is a major cause of ischemic stroke and vascular dementia, which are among the leading causes of death and disability worldwide. Metabolomics can help identify novel risk factors to better understand pathogenesis, predict disease progression and severity, and identify therapeutic targets.

**Methods:** We analyzed metabolomics profiles from 118,021 UK Biobank participants with baseline metabolomics measurements (baseline surveys, 2006-2010; latest follow-up, March 2022). We examined cross-sectional associations of 325 metabolites with clinical diagnoses of stroke and dementia and MRI markers of small vessel disease. We also evaluated relationships between metabolites and future risk of stroke and dementia and performed Mendelian randomization to ascertain causal relationships.

**Results:** Among 118,021 participants (54% women; mean recruitment age, 56.5 years), 2,477 stroke and 1,785 dementia events were recorded (median follow-up, 13.1 years). In cross-sectional analyses, lower levels of apolipoproteins, free cholesterol, cholesteryl esters, fatty acids, lipoprotein particle concentrations, phospholipids, and triglycerides were associated with increased white matter microstructural damage on diffusion tensor MRI. Lower levels of amino acids and fatty acids and higher levels of ketone bodies were associated with increased risk of dementia. In longitudinal analyses, lipoprotein subclasses of very large HDL were associated with increased risk of stroke, and acetate, 3-hydroxybutyrate, and relative lipoprotein lipid concentrations were associated with increased risk of dementia. Mendelian randomization analyses identified strong evidence supporting causal relationships for many associations.

**Conclusions:** biomarker profiling platform and experimentation In this large-scale metabolomics study, we found multiple metabolites associated with stroke, dementia, and MRI markers of small vessel disease. Further studies may help develop personalized prediction models for patients at increased risk of stroke and dementia and provide insights into mechanistic pathways and future treatment approaches.

**Key Points:** *Question:* Are metabolites associated with stroke, dementia, and MRI markers of cerebral small vessel disease?

*Findings:* In an analysis of individual-participant data from 118,021 participants, we identified 289 metabolites that were significantly associated with stroke, dementia, and MRI markers of small vessel disease.

*Meaning:* Metabolic markers were associated with risk of stroke, dementia, and MRI markers of small vessel disease, which could be used to develop personalized prediction models and novel treatment approaches for patients at increased risk of vascular-related conditions.

## Introduction

There are 6.3 million deaths due to stroke and 1.9 million deaths due to dementia each year,^1^ and 50 million people currently living with dementia.^2^ Improved techniques are needed to identify better predictive markers and uncover novel disease mechanisms to develop new treatments.

Increasing evidence implicates vascular risk factors and chronic cerebrovascular disease in the pathogenesis of not only vascular dementia, but also neurodegenerative dementias such as Alzheimer’s disease (AD).^3^ Recent data have suggested treating vascular risk factors, particularly hypertension, may reduce dementia risk.^4^ This emphasizes the potential for targeting cerebrovascular disease to reduce the burden of all types of dementia. Even with small treatment effects this could have a major global impact.

One specific subtype of cerebrovascular disease which appears closely linked to dementia risk is cerebral small vessel disease (SVD), a disease of the small perforating vessels within the white matter and deep grey matter nuclei. SVD results in lacunar infarcts and more chronic changes seen on MRI, including white matter hyperintensities (WMH), cerebral microbleeds, enlarged perivascular spaces, brain atrophy, and diffuse white matter damage identified using diffusion tensor imaging (DTI).^5^ SVD is the major pathology underlying vascular dementia, and it also interacts with Alzheimer’s pathology to increase the probability of developing clinical dementia.^6^ Therefore it represents an important treatment target not only to reduce vascular dementia, but also to reduce the impact of neurodegenerative dementias such as AD.

Metabolomics, the high-throughput identification and quantification of small molecules in biological samples,^7^ enables detailed quantification of metabolic phenotypes, which can be used to identify novel biomarkers to diagnose and monitor disease and characterize metabolic pathways underlying disease pathogenesis.^8^ Metabolomics has been applied in cardiovascular and dementia research previously,^9–11^ but most of the studies have been relatively small (less than 10,000 individuals). In this analysis we analyzed metabolomics profiles from 118,021 participants in the UK Biobank to characterize cross-sectional and longitudinal associations of 325 metabolites with clinical diagnoses of stroke and dementia and MRI markers of SVD. We also ascertained whether these relationships are likely to be causal using Mendelian randomization.

## Methods

### Data source

UK Biobank is a prospective cohort study of over 500,000 participants recruited from 22 centers across the United Kingdom.^12^ Participants were aged between 40-69 at baseline assessment in 2006-2010. Participants completed a comprehensive questionnaire and verbal interview and provided blood samples for metabolomics assays and genetic analyses.

### Metabolomics data

Metabolic biomarker profiling of EDTA plasma samples was performed at Nightingale Health’s laboratories in Finland from a random subset of 121,695 non-fasting participants, of which 118,021 were from baseline recruitment (2006-2010).^13^ High-throughput nuclear magnetic resonance (NMR) spectroscopy was used to obtain 249 metabolic measures, 168 in absolute levels and 81 ratio measures, covering both routine biomarkers and emerging biomarkers with medical relevance (**Supplementary Table 1**). The biomarkers included detailed measures of cholesterol metabolism, fatty acid compositions, and various low-molecular weight metabolites, such as amino acids, ketones, and glycolysis metabolites. For 14 lipoprotein subclasses, the lipid concentrations and composition were measured in terms of triglycerides, phospholipids, total cholesterol, cholesterol esters, free cholesterol, and total lipid concentration within each subclass.

The samples were prepared directly in 96 well-plates by UK Biobank. At least 85 μL plasma was aliquoted into each well using TECAN freedom EVO 150 robotic liquid handlers, which have coefficients of variation in pipetting volume at <0.75% across 8 tips. Plasma samples were shipped to Nightingale Health’s laboratories on dry ice in sample batches of ~5,000-20,000.

Details of the metabolic biomarker profiling platform and experimentation have been described previously.^14,15^ In brief, EDTA plasma samples were stored at −80°C. Before preparation, frozen samples were slowly thawed at +4°C overnight, and then mixed gently and centrifuged (3 min, 3400’g, +4°C) to remove possible precipitate. Aliquots of each sample were transferred into 3-mm outer-diameter NMR tubes and mixed in a 1:1 ratio with a phosphate buffer (75mM Na_2_HPO_4_ in 80%/20% H_2_O/D_2_O, pH 7.4, including also 0.08% sodium 3-(trimethylsilyl) propionate-2,2,3,3-d_4_ and 0.04% sodium azide) automatically with an automated liquid handler (PerkinElmer Janus Automated Workstation).

The prepared samples were loaded onto a cooled sample changer, which maintained the temperature of samples waiting to be measured at +6°C. Two spectra were recorded for each plasma sample using a 500 MHz NMR spectrometer (Bruker AVANCE IIIHD). The first spectrum was a presaturated proton NMR spectrum, which featured resonances arising mainly from proteins and lipids within various lipoprotein particles. The other spectrum was a T2-relaxation-filtered spectrum where most of the broad macromolecule and lipoprotein lipid signals were suppressed, leading to enhanced detection of low-molecular-weight metabolites. Automated quality control of the spectral data was performed. The metabolic biomarkers were quantified using Nightingale Health’s proprietary software (Nightingale Health biomarker quantification library 2020).

To account for technical variation in metabolite levels, a multi-step processing procedure was applied as described previously.^16^ In brief, for the 168 biomarkers that were measured in absolute levels, the concentrations were log transformed and adjusted in a robust linear regression model that accounted for the time between sample preparation and sample measurement, systematic differences between rows and columns on the 96-well shipment plates, and drift over time within each of the six spectrometers. The 81 composite biomarkers and biomarker ratios provided by UK Biobank were recomputed from their adjusted parts, and an additional 76 biomarker ratios of potential biological significance were computed, resulting in a total of 325 metabolites (**Supplementary Table 1**).

### Clinical and imaging endpoints

In the full set of UK Biobank participants with metabolomics data at baseline (n=118,021), we examined clinical endpoints for all stroke, ischemic stroke, intracerebral hemorrhage, all-cause dementia, Alzheimer’s disease, and vascular dementia. Incident stroke and stroke subtypes and all-cause dementia were defined based on the earliest recorded date that the outcome occurred after baseline assessment, either from self-report or linked hospital admission electronic health records (EHR) in the primary or secondary position, or linked death register records in the underlying cause or any other position. Incident Alzheimer’s disease and vascular dementia were based on linked EHR or death register records only. Identification of linked hospital admission EHR and death register records for each endpoint was based on corresponding ICD-9 or ICD-10 codes (**Supplementary Table 2**).

In participants with metabolomics data for whom MRI had been performed (n=10,024), we examined white matter hyperintensities (WMH) volume and several DTI metrics of white matter tracts. For WMH volume we used a UK Biobank-derived phenotype, the total volume of WMH from T1 and T2 FLAIR images (measured in cubic millimeters),^17^ which we log-transformed for analysis. To obtain DTI metrics, we performed principal component analyses on UK Biobank-derived variables for 48 markers of both mean diffusivity (MD, the degree of diffusion) and fractional anisotropy (FA, the directionality of diffusion) on the FA skeleton of the diffusion brain MRI data, and selected the first principal component of each as summary measures of MD and FA.^18^ From the original DTI-MRI scans we also derived peak width of skeletonized mean diffusivity (PSMD, an automated measure based on skeletonization and histogram analysis.^19^ To obtain comparable effect sizes across outcomes, values for the four imaging markers were rescaled using mean-centering and dividing by the standard deviation across participants.

### Cross-sectional and longitudinal analyses

We performed cross-sectional analyses examining the association of clinical and MRI endpoints per 1-SD higher metabolite levels. We constructed linear regression models for continuous outcomes and logistic regression models for binary outcomes with adjustment for age at recruitment and sex. We also constructed regression models with adjustment for a wide range of possible confounders and vascular risk factors. These analyses were adjusted for age at recruitment, sex, UK Biobank recruitment center, NMR spectrometer, Townsend deprivation index at recruitment, taking blood pressure medications or statins at recruitment, body mass index at recruitment, smoking status at recruitment, and type 2 diabetes mellitus status (based on verbal interview, touchscreen self-report, or linked EHR or death register records).

We also performed longitudinal analyses to determine whether metabolites measured at baseline predicted long-term progression to stroke and dementia, for which we constructed Cox proportional-hazards regression models adjusted for age at recruitment and sex to assess the association of progression to stroke and dementia per 1-SD higher metabolite levels. Again, we also conducted these analyses with adjustment for the possible confounders and vascular risk factors listed above.

### Mendelian randomization analyses

To assess whether the association of metabolites with stroke and dementia were causal, we performed two-sample Mendelian randomization, which uses genetic variants as instrumental variables in an approach analogous to a randomized controlled trial.^20^ We obtained summary statistics from genome-wide association studies of each metabolite measured in 115,078 participants from UK Biobank, conducted by the MRC Integrative Epidemiology Unit at Bristol.^21^ Summary statistics for late-onset Alzheimer’s disease were obtained from the International Genomics of Alzheimer’s Project (IGAP).^22^ We obtained summary statistics for lacunar stroke from a previously published genome-wide association study involving 7,338 cases and 254,798 controls.^23^ Summary statistics for stroke and ischemic stroke subtypes were obtained from the MEGASTROKE Consortium,^24^ consisting of 67,162 cases and 454,450 controls, which we restricted to Europeans. There were 60,341 cases of ischemic stroke, 9,006 cases of cardioembolic stroke, and 6,688 cases of large artery stroke. Summary statistics for WMH (n=42,310), MD (n=17,467), and FA (n=17,663) were obtained from a genome-wide association study of participants from UK Biobank and the CHARGE Consortium.^25^ Summary statistics for PSMD were obtained from a currently unpublished genome-wide association study. Our primary Mendelian randomization analyses used inverse-variance weighted meta-analysis under a random effects model (to account for heterogeneity) to combine the ratio estimates from each genetic variant into a single estimate of the causal effect of each metabolite on each outcome.^20^ We conducted sensitivity analyses using a variety of robust Mendelian randomization methods, which employ different assumptions to make reliable causal inferences. These included MR-Egger regression, weighted median estimator, and simple and weighted mode-based estimators. For each metabolite, we harmonized all SNPs associated with that metabolite with the outcome data to ensure that effect estimates of each SNP on the metabolite and outcome corresponded to the same effect allele. We then performed Mendelian randomization using the IVW method and additional methods as sensitivity analyses.

All statistical analyses were conducted using R version 4.1.1 (R Core Team, 2021). To account for multiple testing comparisons, we used a false discovery rate (FDR) threshold of *q* < 0.05 to identify significant associations for each outcome measure. Two-sided *P*-values and 95% confidence intervals are presented. Mendelian randomization analyses were conducted using the TwoSampleMR package version 0.5.4.

## Results

### Participant characteristics

We analyzed data from 118,021 participants from UK Biobank with metabolomics measurements, of whom 54% were female and 95% were white, with a mean age of 56.5 (SD: 8.1) years (**Table 1**). The median follow-up time was 13.1 years (5^th^-95^th^ percentile, 11.8-14.4 years).

**Table 1.** Characteristics of UK Biobank participants with metabolomics data at baseline

| Trait | Mean (SD) or n (%) | N |
| --- | --- | --- |
| **Risk factors** |  |  |
| Female sex | 63,890 (54.2%) | 117,938 |
| Age at recruitment (years) | 56.5 (8.1) | 118,021 |
| Ethnicity (White) | 111,323 (94.8%) | 117,491 |
| Townsend deprivation index at recruitment | -1.31 (3.10) | 117,864 |
| Body mass index (BMI) (kg/m<sup>2</sup>) | 27.43 (4.78) | 117,609 |
| Systolic blood pressure (mm Hg) | 137.8 (18.6) | 117,902 |
| Diastolic blood pressure (mm Hg) | 82.2 (10.2) | 117,903 |
| Current smoker | 12,474 (10.6%) | 117,449 |
| Has Type 2 diabetes mellitus | 11,277 (9.6%) | 118,020 |
| Has hypertension | 53,999 (45.8%) | 117,902 |
| Taking blood pressure medication | 24,305 (20.8%) | 117,079 |
| Taking statins | 21,199 (24.1%) | 88,067 |
| APOE ε4 carrier | 29,628 (25.1%) | 118,021 |
| **Outcomes** |  |  |
| Log white matter hyperintensities (WMH) volume | 8.02 (1.01) | 9,998 |
| Fractional anisotropy (FA) | 0.0085 (4.35) | 9,779 |
| Mean diffusivity (MD) | 0.0379 (4.44) | 9,779 |
| Peak width of skeletonized mean diffusivity (PSMD) | 2.28 x 10<sup>-4</sup> (3.97 x 10<sup>-5</sup>) | 9,710 |
| Incident all stroke case | 2,477 (2.1%) | 118,021 |
| Incident ischemic stroke case | 2,053 (1.7%) | 118,021 |
| Incident intracerebral hemorrhage case | 464 (0.4%) | 118,021 |
| Incident all-cause dementia case | 1,785 (1.5%) | 118,021 |
| Incident Alzheimer's disease case | 765 (0.6%) | 118,021 |
| Incident vascular dementia case | 418 (0.3%) | 118,021 |

### Associations with baseline imaging parameters

We analyzed the association of 325 metabolic measures with clinical endpoints and MRI markers in cross-sectional analyses adjusted for age at recruitment and sex (**Supplementary Figure 1, Supplementary Table 3**). We found that lower levels of apolipoproteins, cholesterol, free cholesterol, cholesteryl esters, fatty acids, lipoprotein particle concentrations, phospholipids, triglycerides, and total lipids, and higher levels of amino acids, glucose, and glycoprotein acetyls (an inflammatory marker) were associated with increased white matter microstructural damage on DTI, as indicated by higher WMH, MD, and PSMD, and lower FA. These metabolites were also associated with increased risk of all stroke, ischemic stroke, all-cause dementia, and vascular dementia.

In cross-sectional analyses adjusted for possible confounders and additional vascular risk factors (age at recruitment, sex, UK Biobank recruitment center, spectrometer, Townsend deprivation index, blood pressure medication, statins, body mass index, smoking status, and diabetes status), we found many of the associations with stroke and dementia attenuated and were no longer significant. However, most of the metabolites remained significantly associated with FA, MD, and PSMD (**Figure 1, Supplementary Table 4**). Lower levels of apolipoproteins, cholesterol, free cholesterol, cholesteryl esters, fatty acids, lipoprotein particle concentrations, phospholipids, triglycerides, and total lipids were associated with higher MD and PSMD and lower FA. Additionally, lower levels of albumin and higher levels of creatinine and glucose were associated with increased risk of all stroke and ischemic stroke, while lower levels of amino acids and higher levels of ketone bodies were associated with increased risk of all-cause dementia.

**Figure 1.**
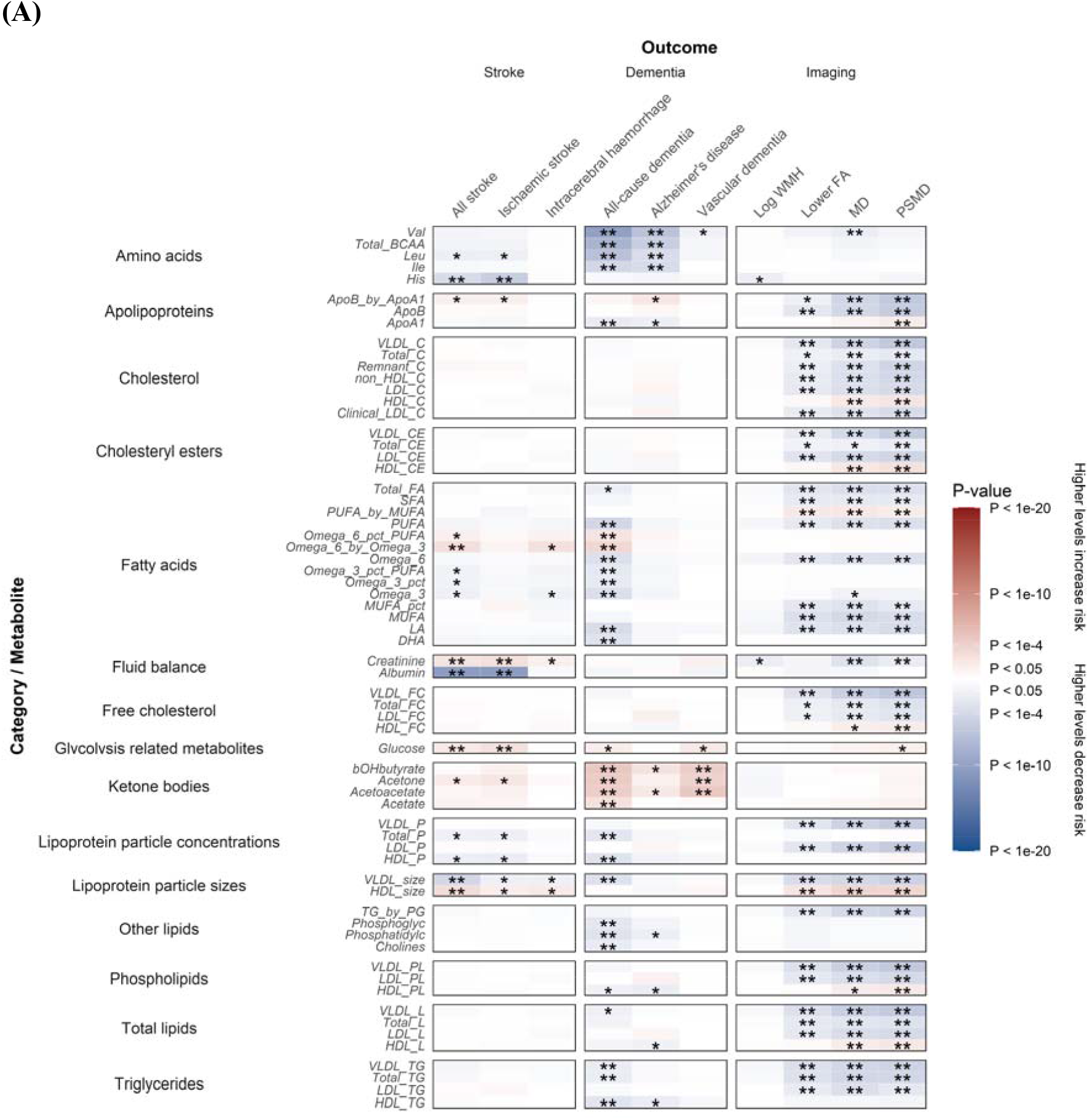
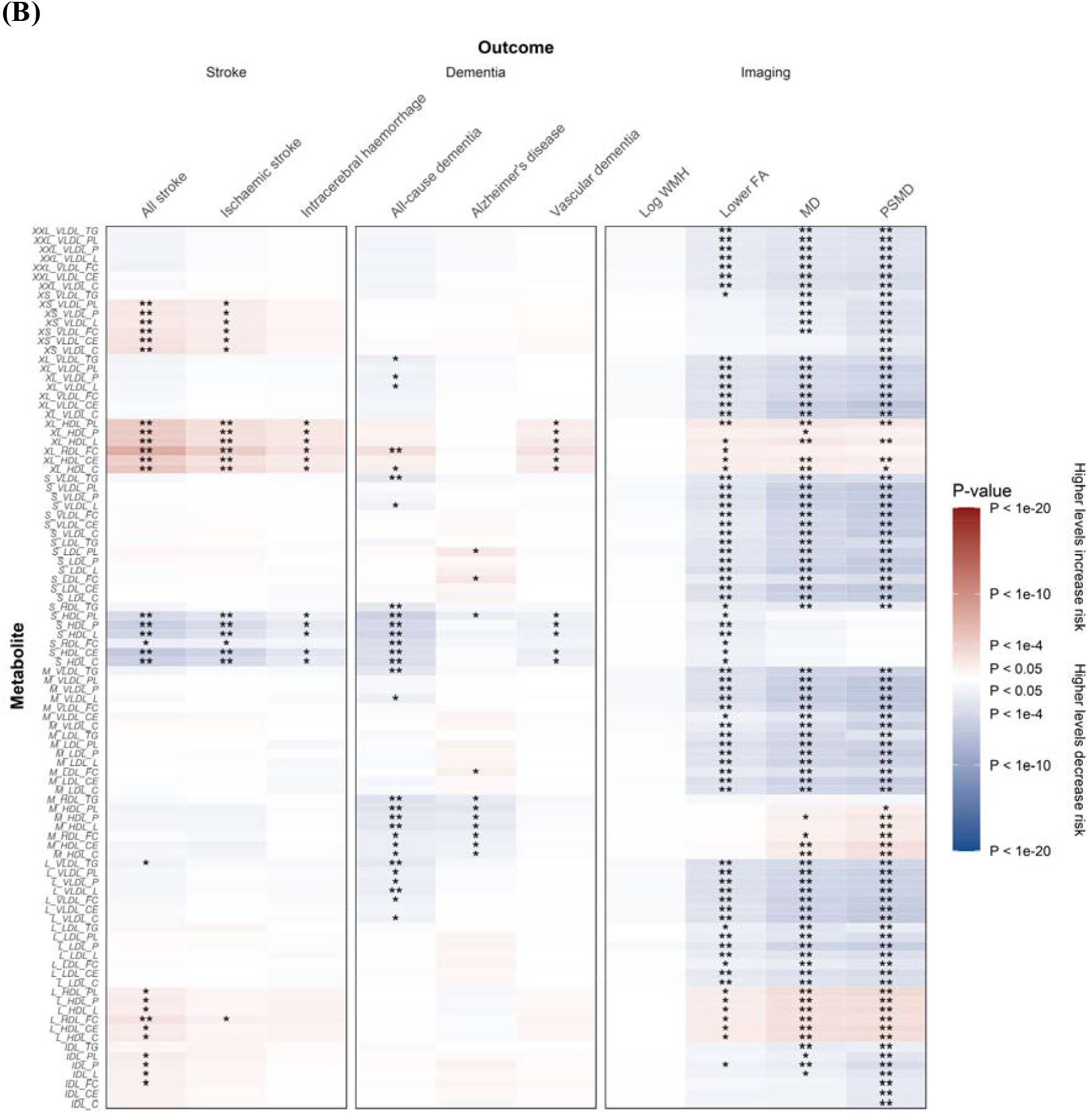
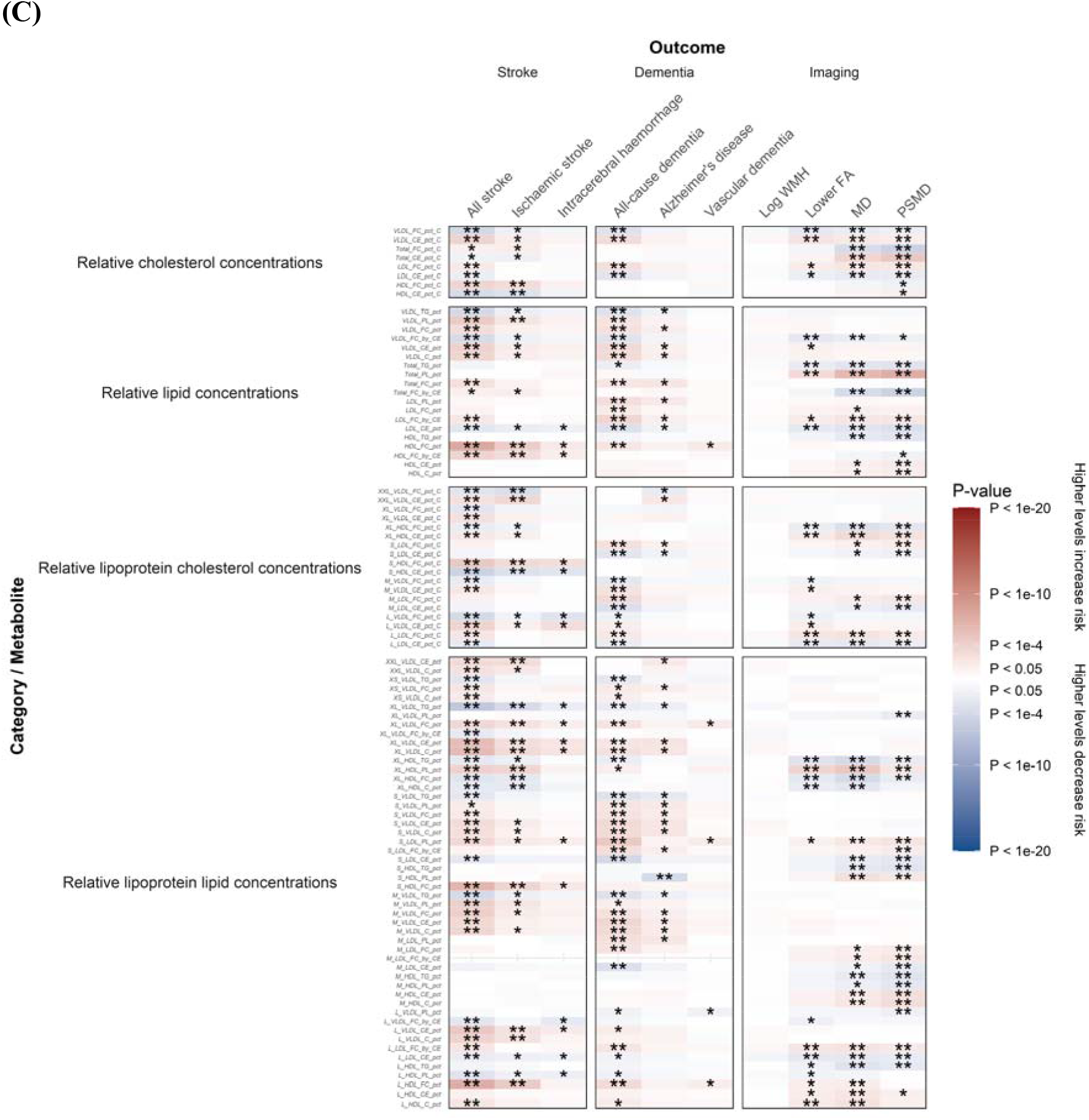
Association of stroke, dementia, and MRI markers at baseline per 1-SD higher metabolite levels with adjustment for possible confounders and vascular risk factors. (**A**) Lipids and other metabolites. (**B**) Lipoprotein subclasses. (**C**) Relative lipid, lipoprotein, and cholesterol concentrations. Beta estimates and *P*-values were obtained from linear or logistic regression models adjusted for age at recruitment, sex, UK Biobank recruitment center, Townsend deprivation index at recruitment, whether the person was taking blood pressure medication or statins, body mass index, smoking status, and Type 2 diabetes mellitus status. Colors show magnitude and direction of *P*-value for association of metabolite with each outcome (red indicates positive association and blue indicates inverse association). Asterisks indicate significance: \**P* < 0.05; **FDR *q* < 0.05.

### Longitudinal analyses of stroke and dementia

When accounting for long-term follow-up in time-to-event analyses adjusted for age and sex (**Supplementary Figure 1, Supplementary Table 5**), higher levels of the ratio of free cholesterol to cholesteryl esters in very small VLDL and higher levels of glycoprotein acetyls were associated with increased risk of all stroke. Conversely, higher levels of docosahexaenoic acid (DHA) and several lipoprotein subclasses containing cholesterol, free cholesterol, and cholesteryl esters were associated with decreased risk of all stroke. Higher levels of glucose and the percentage of phospholipids to total lipids in very small VLDL were associated with increased risk of all-cause dementia, while a wide range of cholesterol, cholesteryl esters, free cholesterol, apolipoprotein B, and lipoprotein subclasses were associated with decreased risk of all-cause dementia.

In longitudinal analyses adjusted for potential confounders and vascular risk factors (**Figure 2, Supplementary Table 6**), total lipids in very large HDL were most strongly associated with all stroke. For all-cause dementia, acetate, 3-hydroxybutyrate, and the ratio of free cholesterol to cholesteryl esters in small LDL were associated with increased risk of dementia. Meanwhile, omega-3 fatty acids, DHA, leucine, isoleucine, valine, the total concentration of branched-chain amino acids, and several lipoprotein subclasses were associated with decreased risk of dementia.

**Figure 2.**
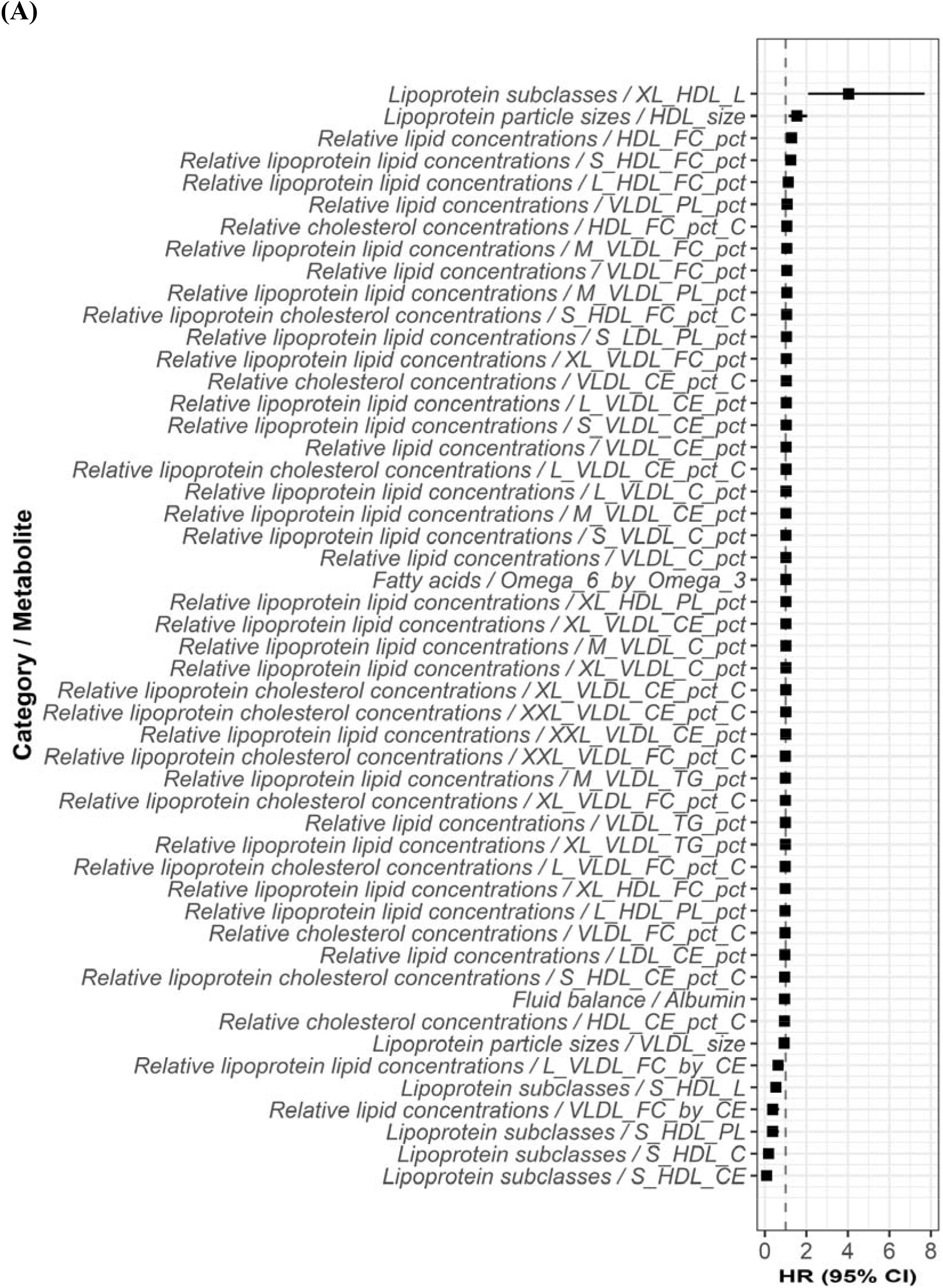
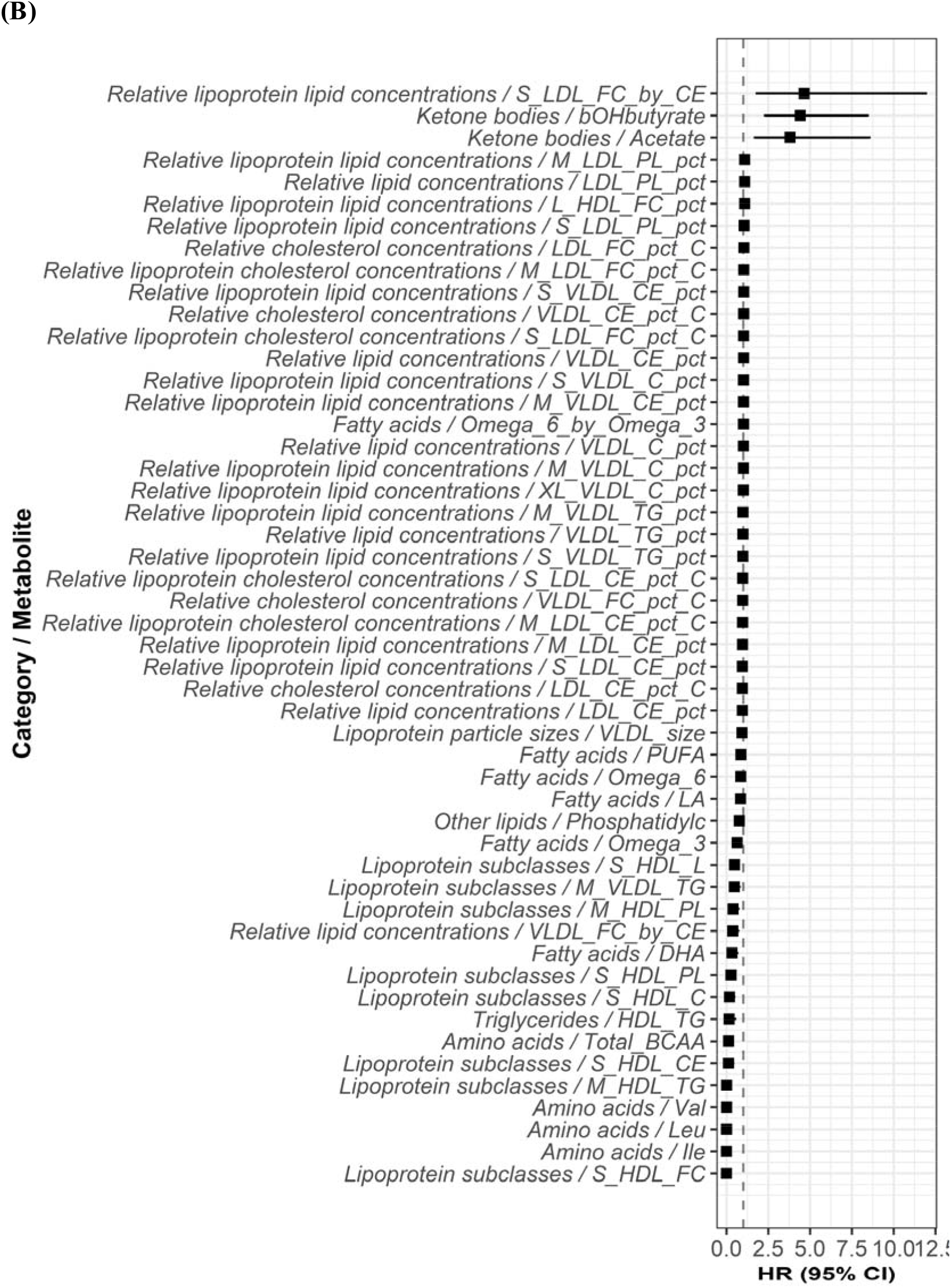
Adjusted hazard ratios for all stroke and all-cause dementia per 1-SD higher metabolite levels with adjustment for possible confounders and vascular risk factors. **(A)** All stroke. **(B)** All-cause dementia. Analyses were adjusted for age at recruitment and sex. Filled squares indicate associations significant at FDR *q* < 0.05.

### Mendelian randomization analyses

Using Mendelian randomization, we found genetically elevated levels of cholesteryl esters in very large VLDL and total lipids in small VLDL were associated with increased risk of ischemic stroke, and genetically lowered levels of the concentration of HDL particles were associated with increased risk of lacunar stroke (**Figure 3, Supplementary Table 7**). Furthermore, genetically elevated levels of LDL within cholesterol, cholesteryl esters, free cholesterol, and phospholipids, and lipoprotein subclasses of LDL, VLDL, and IDL, and genetically lower levels of total lipids in medium HDL, were associated with increased risk of late-onset AD.

**Figure 3.**
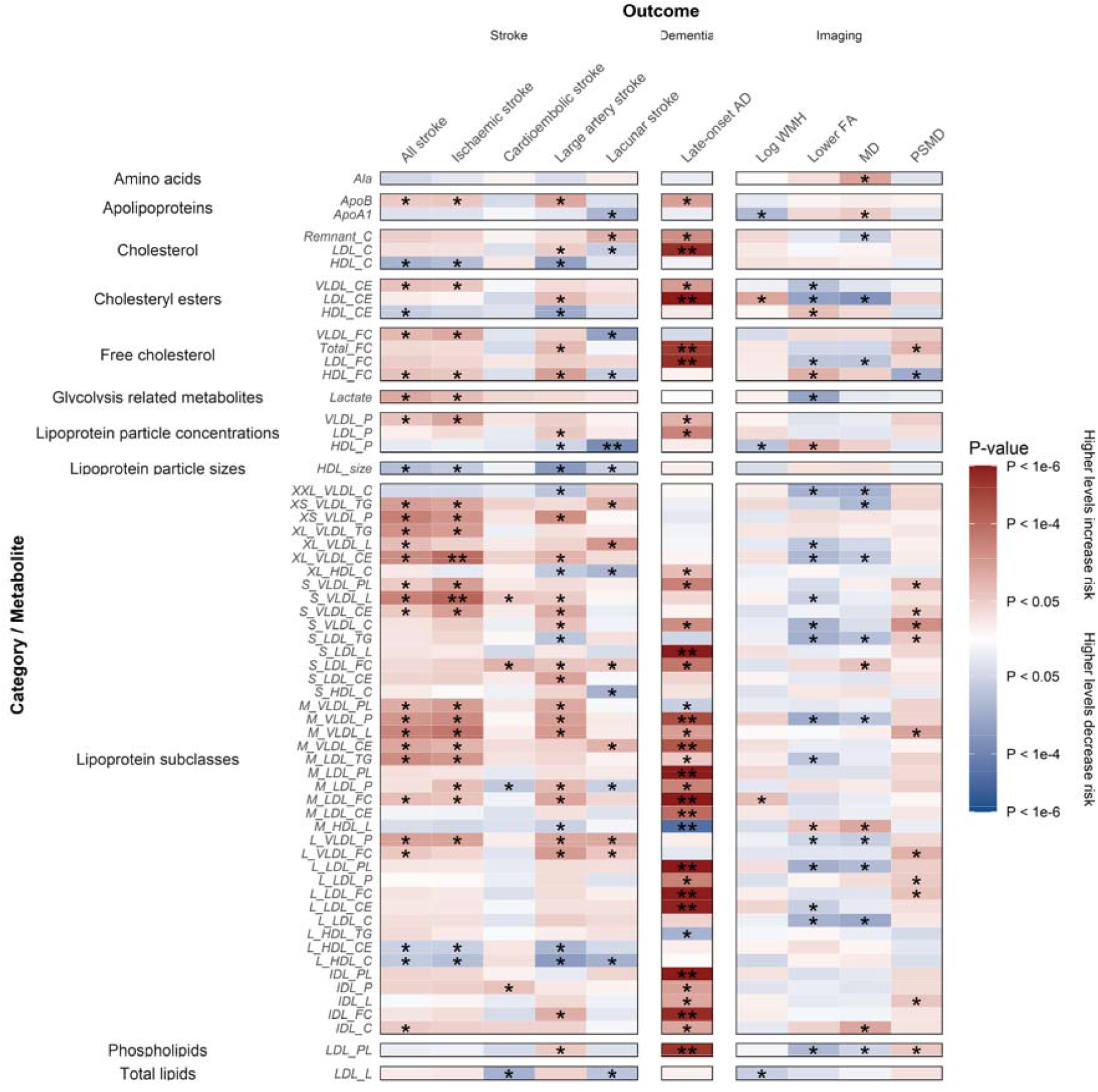
Mendelian randomization results showing causal estimates for association of metabolite levels with stroke, dementia, and MRI markers. Colors show magnitude and direction of *P*-value of association for estimate of causal effect using inverse-variance weighted Mendelian randomization approach (red indicates positive association and blue indicates inverse association). Asterisks indicate significance: \**P* < 0.05; **FDR *q* < 0.05.

## Discussion

In this large-scale metabolomics study of 118,021 individuals, we identified 289 metabolic markers that are significantly associated with stroke, dementia, and MRI markers of SVD. We found that lower levels of apolipoproteins, cholesterol, free cholesterol, cholesteryl esters, fatty acids, lipoprotein particle concentrations, phospholipids, triglycerides, and total lipids were associated with increased white matter microstructural damage. Additionally, lower levels of amino acids and fatty acids and higher levels of ketone bodies were associated with increased risk of all-cause dementia. Increased levels of lipoprotein subclasses of large HDL, very large HDL, and very small VLDL were associated with increased risk of stroke and dementia, whereas lipoprotein subclasses of small and medium HDL were associated with decreased risk of stroke and dementia.

Although there is a well-established inverse association of plasma HDL cholesterol levels with coronary heart disease, stroke, and vascular brain damage, whether these relationships are causal has remained uncertain.^26–33^ This study provides new insights on these relationships by demonstrating that the direction and magnitude of the association of HDL with stroke and dementia and their subtypes depends on the size of the lipoprotein subclasses within HDL.

The metabolite associations that we observed for all-cause dementia, AD, and vascular dementia confirm the findings reported in a recent preprint.^34^ However, we analyzed an expanded set of metabolites by including 76 additionally derived biomarkers of potential biological significance, and evaluated associations with a wider range of endpoints including stroke, ischemic stroke, intracerebral hemorrhage, and DTI markers.

Our findings have several important clinical implications. First, they may provide novel insights into the metabolic pathways underlying stroke and dementia. It is possible that modifying levels of specific metabolites could help reduce the risk of vascular-related conditions, so this research could help inform dietary interventions or the development of novel therapies. Second, a metabolomics panel based on these associations could be developed for clinicians to predict which patients are most likely to develop stroke and dementia and offer personalized treatment plans.

Our study has several strengths. First, it is one of the largest metabolomics studies conducted to date. Having metabolomics data available in nearly 120,000 individuals greatly increases the power to detect statistically significant associations. Second, the metabolites were measured using a fully automated, comprehensive spectrum analysis under strict quality control, which increases the accuracy and validity of the findings. Third, the prospective study design and long follow-up period, with metabolites that were measured prior to disease onset, was particularly useful for evaluating their associations with risk of stroke and dementia. Fourth, we conducted sensitivity analyses to assess the impact of potential confounders and vascular risk factors on the identified associations.

Our study also has limitations. First, we conducted a large number of statistical tests, so we applied an FDR threshold to reduce the likelihood of identifying false positives. However, some associations may have been biologically and clinically meaningful but did not reach the statistical significance threshold after correction for multiple testing. Second, the study sample was large but is not representative of the wider UK population. Third, in UK Biobank we only had endpoints of ischemic and hemorrhagic stroke but not subtypes of ischemic stroke (e.g. large artery stroke, cardioembolic stroke, and lacunar stroke). This meant that we were unable to examine associations of metabolites directly with lacunar stroke, which one might expect if metabolites were associated with MRI markers of SVD. Nevertheless, our Mendelian randomization analyses identified evidence that genetically elevated levels of metabolites are associated with increased risk of lacunar stroke. Fourth, we obtained summary statistics of genetic associations with late-onset AD but not with all-cause dementia and vascular dementia, which limited the scope of our assessment of causal relationships of metabolites with dementia.

## Conclusions

We found evidence supporting the association of a wide range of metabolites with stroke, dementia, and MRI markers of SVD. Although further research is needed, these findings could be used to help develop personalized prediction models and novel treatment approaches.

## Supporting information

STROBE Checklist

## Data Availability

Researchers can apply to access the UK Biobank raw data at https://www.ukbiobank.ac.uk/enable-your-research. All data produced in the present work are contained in the manuscript and its supplementary materials.

## Acknowledgements

The authors thank Daniel Tozer (Department of Clinical Neurosciences, University of Cambridge) for his assistance with the derivation of PSMD.

## Author Contributions

ELH and HSM had full access to all the data in the study and take responsibility for the integrity of the data and the accuracy of the data analysis. *Concept and design:* ELH and HSM. *Acquisition, analysis, or interpretation of data:* ELH and HSM. *Drafting of the manuscript:* ELH and HSM. *Critical revision of the manuscript for important intellectual content:* ELH and HSM. *Statistical analysis:* ELH. *Obtained funding:* ELH and HSM. *Administrative, technical, or material support:* HSM. *Supervision:* HSM.

## Conflict of Interest Disclosures

The authors report nothing to disclose.

## Funding/Support

This research was conducted using the UK Biobank under application number 36509. It was funded by the British Heart Foundation via a fellowship awarded to ELH from the Cambridge BHF Centre of Research Excellence (RE/18/1/34212) and a BHF program grant (RG/4/32218). Infrastructural support was provided by the Cambridge University Hospitals NIHR Biomedical Research Centre (BRC-1215-20014). HSM is supported by a NIHR Senior Investigator Award.

## Role of the Funder/Sponsor

The funders had no role in the design and conduct of the study; collection, management, analysis, and interpretation of the data; preparation, review, or approval of the manuscript; and decision to submit the manuscript for publication.

## Disclaimer

The views expressed in this publication are those of the authors and not necessarily those of the NIHR, NHS, or UK Department of Health and Social Care.

## Notes

### Competing Interest Statement

The authors have declared no competing interest.

### Author Declarations

UK Biobank has approval from the North West Multi-centre Research Ethics Committee (MREC) as a Research Tissue Bank (RTB) approval. This research was conducted using the UK Biobank under application number 36509.

