## Supplementary Figures for "Metabolic associations with stroke, dementia, and imaging markers of cerebral small vessel disease: a comprehensive metabolomics study"

### Supplementary Material

**Supplementary Tables**

Supplementary Table 1. Metabolic measures analyzed using nuclear magnetic resonance spectroscopy in UK Biobank

Supplementary Table 2. International Classification of Diseases codes for endpoints included in analysis

Supplementary Table 3. All results for association of stroke, dementia, and MRI markers at baseline per 1-SD higher metabolite levels with basic adjustment

Supplementary Table 4. All results for association of stroke, dementia, and MRI markers at baseline per 1-SD higher metabolite levels with further adjustment for possible confounders and vascular risk factors

Supplementary Table 5. All results from Cox proportional-hazards regression models for stroke and dementia per 1-SD higher metabolite levels with basic adjustment

Supplementary Table 6. All results from Cox proportional-hazards regression models for stroke and dementia per 1-SD higher metabolite levels with further adjustment for possible confounders and vascular risk factors

Supplementary Table 7. All results from Mendelian randomization analyses of association of metabolite levels with stroke, dementia, and MRI markers

**Supplementary Figures**

Supplementary Figure 1. Association of MRI markers and other outcomes at baseline per 1-SD higher metabolite levels with basic adjustment

Supplementary Figure 2. Adjusted hazard ratios for all stroke and all-cause dementia per 1-SD higher metabolite levels with basic adjustment

**Supplementary Figure 1. Association of stroke, dementia, and MRI markers at baseline per 1-SD higher metabolite levels with basic adjustment.** (**A**) Lipids and other metabolites. (**B**) Lipoprotein subclasses. (**C**) Relative lipid, lipoprotein, and cholesterol concentrations. Beta estimates and *P*-values were obtained from linear or logistic regression models adjusted for age at recruitment and sex. Colors show magnitude and direction of *P*-value for association of metabolite with each outcome (red indicates positive association and blue indicates inverse association). Asterisks indicate significance: **P* < 0.05; **FDR *q* < 0.05.

**(A)**

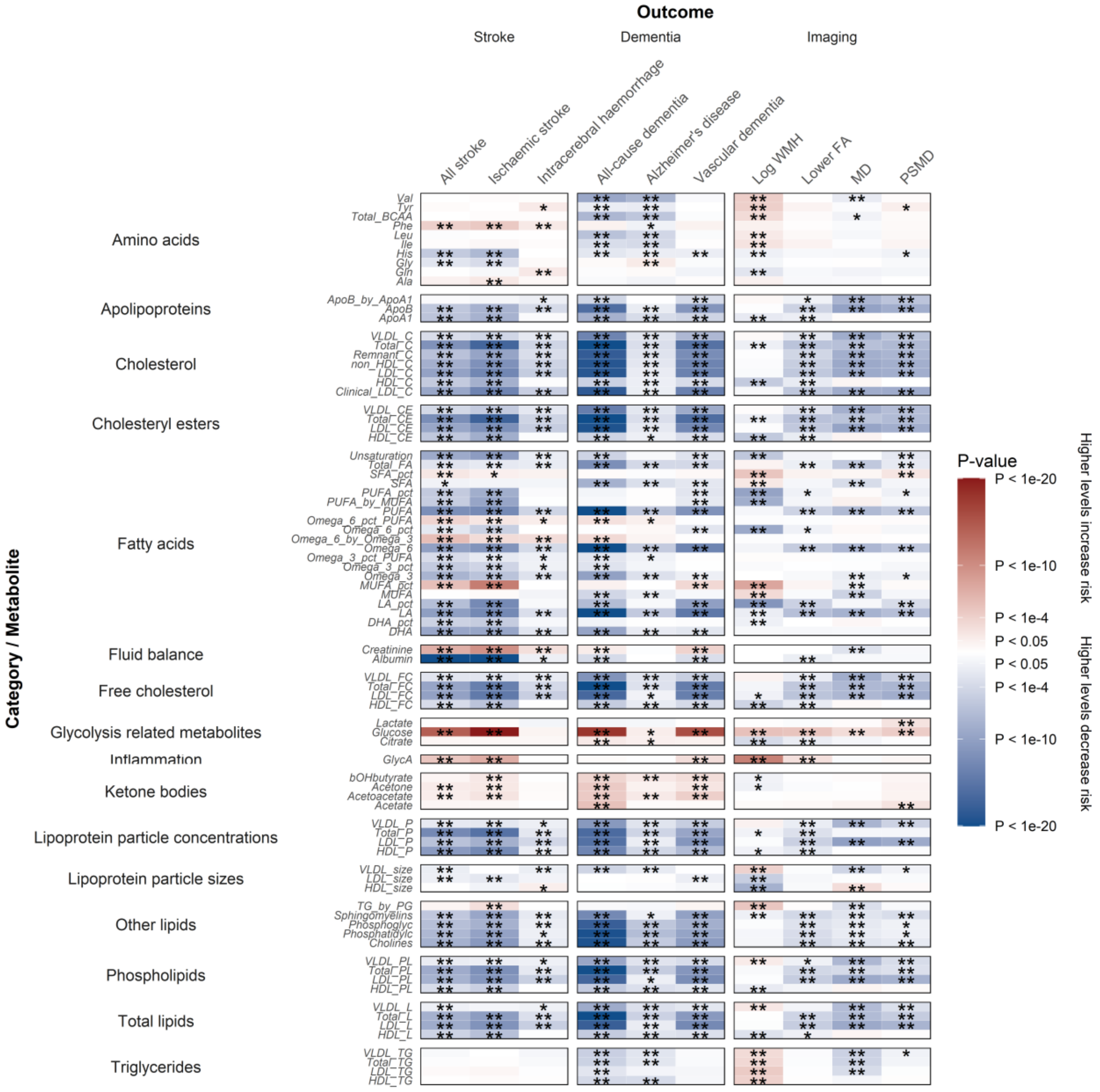

**(B)**

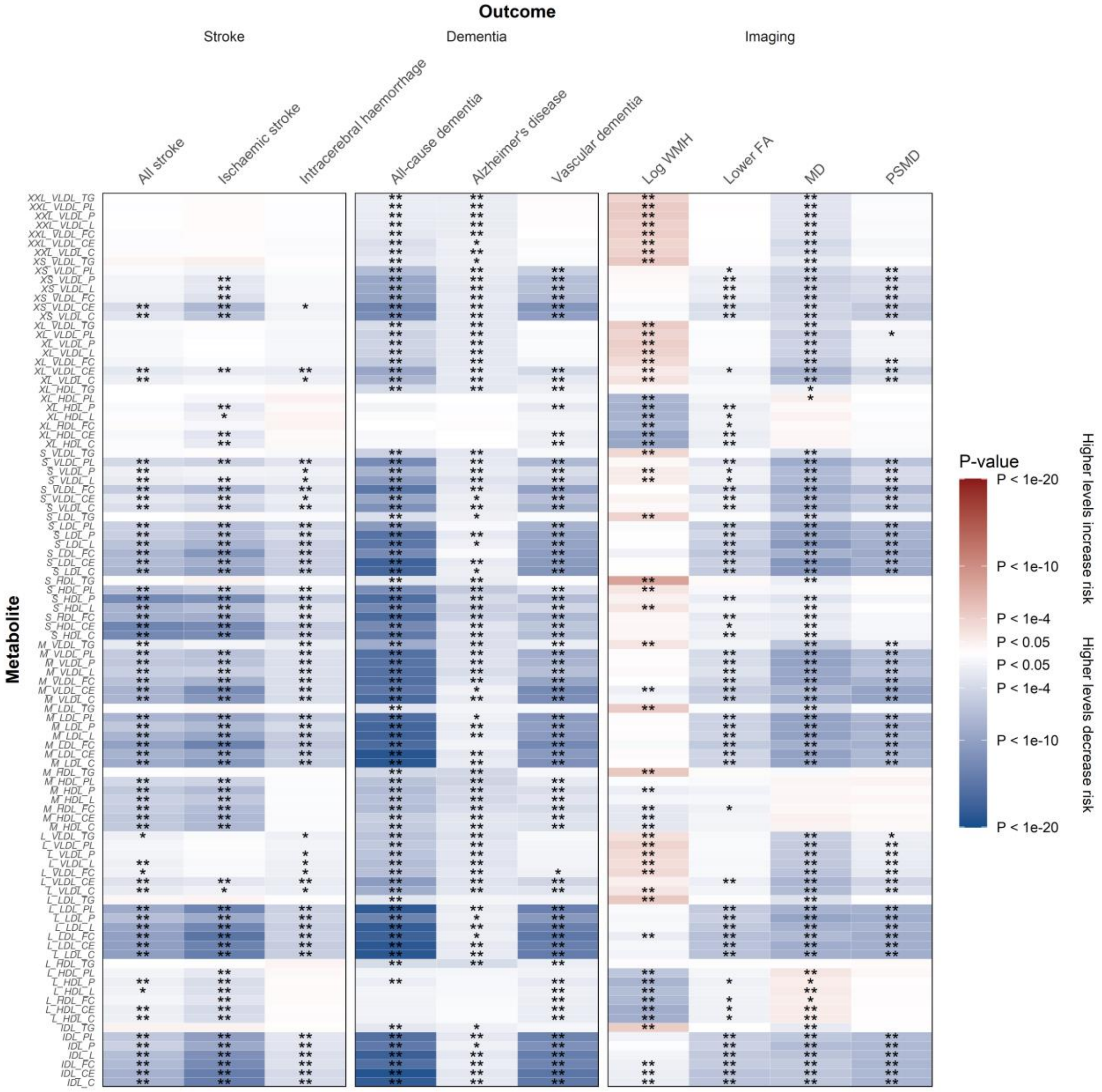
Outcome
Stroke
Dementia
Imaging
All stroke
Ischaemic stroke
Intracerebral haemorrhage
All-cause dementia
Alzheimer's disease
Vascular dementia
Log WMH
Lower FA
MD
PSMD
Metabolite
P-value
P < 1e-20
P < 1e-10
P < 1e-4
P < 0.05
P < 0.05
P < 1e-4
P < 1e-10
P < 1e-20
Higher levels increase risk
Higher levels decrease risk

(C)

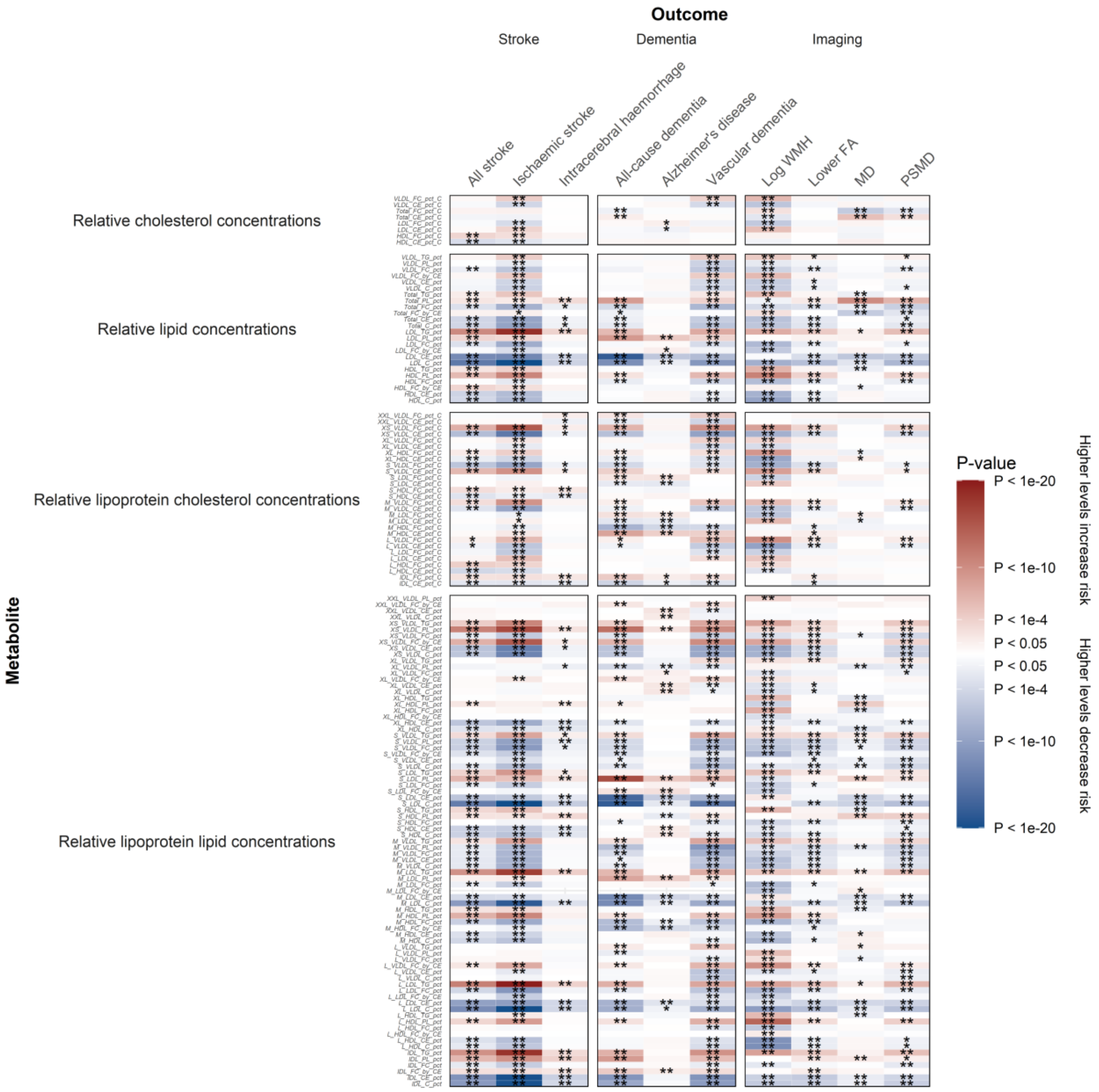

Outcome
Stroke
Dementia
Imaging
All stroke
Ischaemic stroke
Intracerebral haemorrhage
All-cause dementia
Alzheimer's disease
Vascular dementia
Log WMH
Lower FA
MD
PSMD
Metabolite
Relative cholesterol concentrations
Relative lipid concentrations
Relative lipoprotein cholesterol concentrations
Relative lipoprotein lipid concentrations
P-value
P < 1e-20
P < 1e-10
P < 1e-4
P < 0.05
P < 0.05
P < 1e-4
P < 1e-10
P < 1e-20
Higher levels increase risk
Higher levels decrease risk

**Supplementary Figure 2. Adjusted hazard ratios for all stroke and all-cause dementia per 1-SD higher metabolite levels with basic adjustment. (A)** All stroke. **(B)** All-cause dementia. Analyses were adjusted for age at recruitment and sex. Filled squares indicate associations significant at FDR $q < 0.05$.

**(A)**

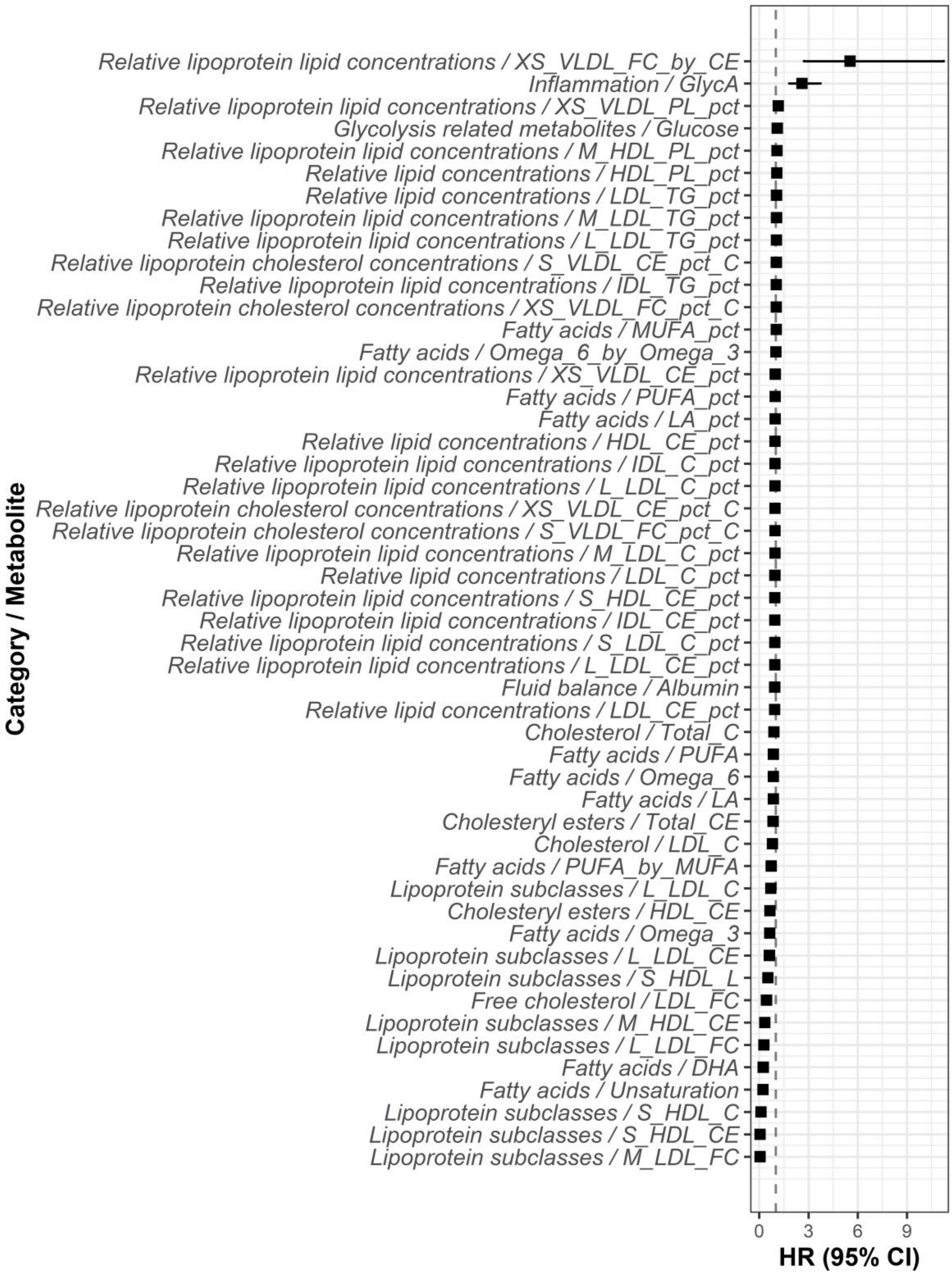

(B)

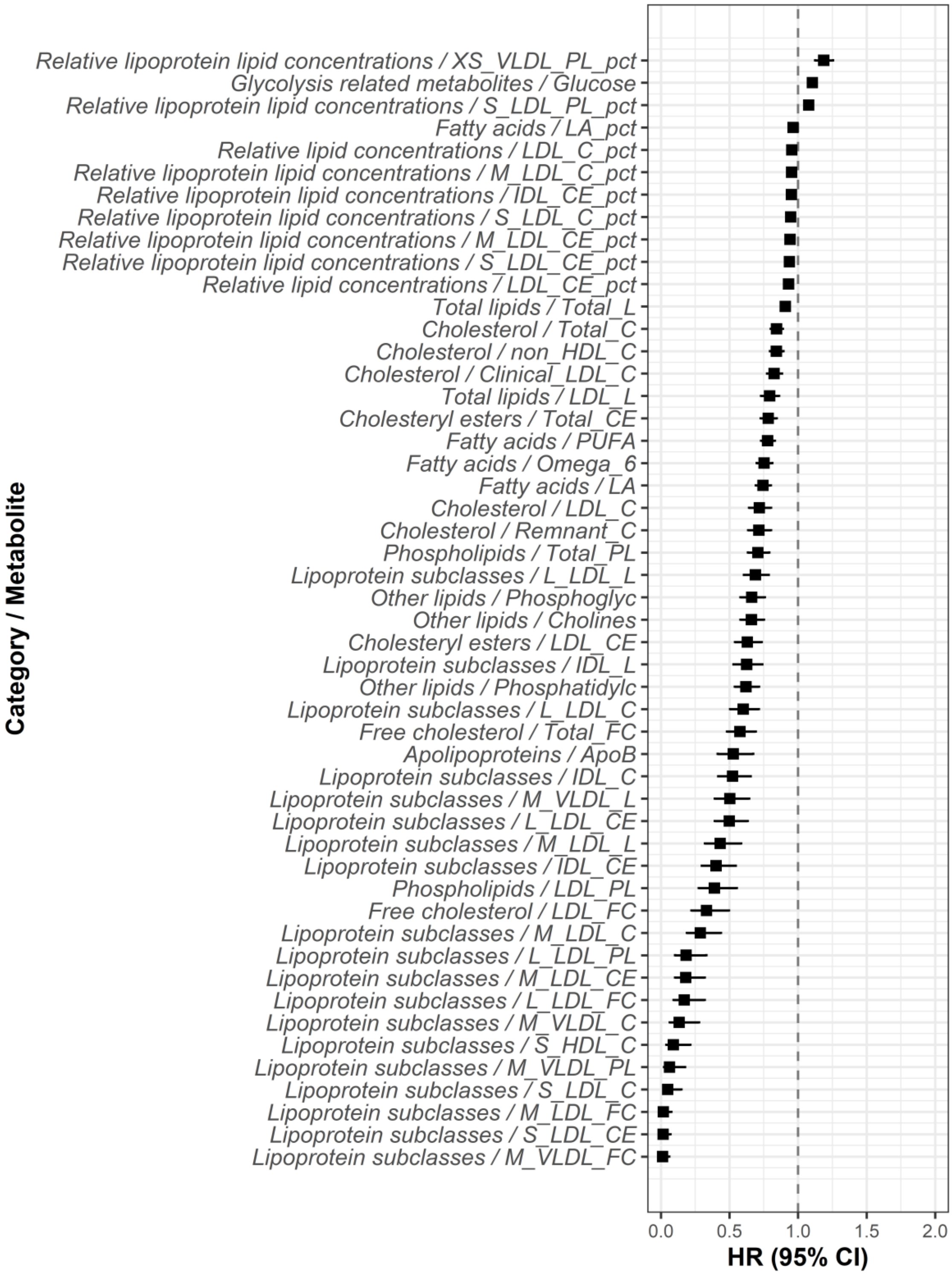
Relative lipoprotein lipid concentrations / XS_VLDL_PL_pct
Glycolysis related metabolites / Glucose
Relative lipoprotein lipid concentrations / S_LDL_PL_pct
Fatty acids / LA_pct
Relative lipid concentrations / LDL_C_pct
Relative lipoprotein lipid concentrations / M_LDL_C_pct
Relative lipoprotein lipid concentrations / IDL_CE_pct
Relative lipoprotein lipid concentrations / S_LDL_C_pct
Relative lipoprotein lipid concentrations / M_LDL_CE_pct
Relative lipoprotein lipid concentrations / S_LDL_CE_pct
Relative lipid concentrations / LDL_CE_pct
Total lipids / Total_L
Cholesterol / Total_C
Cholesterol / non_HDL_C
Cholesterol / Clinical_LDL_C
Total lipids / LDL_L
Cholesteryl esters / Total_CE
Fatty acids / PUFA
Fatty acids / Omega_6
Fatty acids / LA
Cholesterol / LDL_C
Cholesterol / Remnant_C
Phospholipids / Total_PL
Lipoprotein subclasses / L_LDL_L
Other lipids / Phosphoglyc
Other lipids / Cholines
Cholesteryl esters / LDL_CE
Lipoprotein subclasses / IDL_L
Other lipids / Phosphatidylc
Lipoprotein subclasses / L_LDL_C
Free cholesterol / Total_FC
Apolipoproteins / ApoB
Lipoprotein subclasses / IDL_C
Lipoprotein subclasses / M_VLDL_L
Lipoprotein subclasses / L_LDL_CE
Lipoprotein subclasses / M_LDL_L
Lipoprotein subclasses / IDL_CE
Phospholipids / LDL_PL
Free cholesterol / LDL_FC
Lipoprotein subclasses / M_LDL_C
Lipoprotein subclasses / L_LDL_PL
Lipoprotein subclasses / M_LDL_CE
Lipoprotein subclasses / L_LDL_FC
Lipoprotein subclasses / M_VLDL_C
Lipoprotein subclasses / S_HDL_C
Lipoprotein subclasses / M_VLDL_PL
Lipoprotein subclasses / S_LDL_C
Lipoprotein subclasses / M_LDL_FC
Lipoprotein subclasses / S_LDL_CE
Lipoprotein subclasses / M_VLDL_FC
Category / Metabolite
0.0
0.5
1.0
1.5
2.0
HR (95% CI)
